# An integrated genome and phenome-wide association study approach to understanding Alzheimer’s disease predisposition

**DOI:** 10.1101/2022.01.03.22268705

**Authors:** Archita S. Khaire, Courtney E. Wimberly, Eleanor C. Semmes, Jillian H. Hurst, Kyle M. Walsh

## Abstract

**Background:** Genome-wide association studies (GWAS) have identified common, heritable alleles that increase late-onset Alzheimer’s disease (LOAD) risk. We recently published an analytic approach to integrate GWAS and phenome-wide association study (PheWAS) data, enabling identification of candidate traits and trait-associated variants impacting disease risk, and apply it here to LOAD.

**Methods:** PheWAS was performed for 23 known LOAD-associated single nucleotide polymorphisms (SNPs) and 4:1 matched control SNPs using UK Biobank data. Traits enriched for association with LOAD SNPs were ascertained and used to identify trait-associated candidate SNPs to be tested for association with LOAD risk (17,008 cases; 37,154 controls).

**Results:** LOAD-associated SNPs were significantly enriched for associations with 6/778 queried traits, including three platelet traits. The strongest enrichment was for platelet distribution width (PDW) (P=1.2×10^−5^), but no consistent direction of effect was observed between increased PDW and LOAD susceptibility across variants or in Mendelian randomization analysis. Of 384 PDW-associated SNPs identified by prior GWAS, 36 were nominally associated with LOAD risk and 5 survived false-discovery rate correction for multiple testing. Associations confirmed known LOAD risk loci near *PICALM, CD2AP, SPI1*, and *NDUFAF6*, and identified a novel risk locus in the epidermal growth factor receptor (*EGFR*) gene.

**Conclusions:** Through integration of GWAS and PheWAS data, we identify substantial pleiotropy between genetic determinants of LOAD and of platelet morphology, and for the first time implicate EGFR – a mediator of β-amyloid toxicity – in Alzheimer’s disease susceptibility.

## INTRODUCTION

Late-onset Alzheimer’s disease (LOAD) is defined by the development of symptoms after age 65 and is the most common form of dementia. LOAD affects an estimated 5.7 million Americans, a number projected to grow rapidly with demographic trends.(Rabinovici 2019) Genome-wide association studies (GWAS) have greatly enhanced our understanding of the etiology of LOAD.(Bruni, Bernardi, and Gabelli 2020; Lewcock et al. 2020) A recent GWAS meta-analysis identified common heritable risk alleles at 24 statistically independent genetic loci, implicating genes involved in immunity, lipid metabolism, tau and amyloid-β (Aβ) in LOAD pathogenesis.(Kunkle et al. 2019) Although collaborative GWAS meta-analyses including >90k LOAD patients have enabled identification of low-frequency risk alleles, gains in statistical power are increasingly limited. Furthermore, functional genomics studies to reveal the biologic consequences of associated single nucleotide polymorphisms (SNPs) through *in silico* and *in vivo* analyses remain limited.(Andrews, Fulton-Howard, and Goate 2020) The use of polygenic score and Mendelian randomization (MR) approaches,(Shi et al. 2021; Zhang, Wang, and Liu 2021; Li et al. 2021; Handy et al. 2021) gene-pathway analyses,(Gao et al. 2018) and phenome-wide association studies (PheWAS)(Lumsden et al. 2020) can augment traditional GWAS approaches and expand our understanding of the underlying biology behind LOAD pathogenesis.

PheWAS are an approach to analyze a collection of phenotypes for their association with a particular genetic variant of interest, and they have recently been made feasible through the integration of electronic health record (EHR) data with large-scale genomic datasets.(Denny, Bastarache, and Roden 2016) A PheWAS essentially inverts the classic GWAS approach, testing a large collection of phenotypes (rather than a single disease or trait) for association with a single genetic variant (rather than millions of variants distributed across the genome). However, both GWAS and PheWAS typically test SNP-trait associations through the same multivariable regression-based frameworks, including adjustment for subject ancestry to control for population stratification.(Bush, Oetjens, and Crawford 2016)

By expanding the list of phenotypes associated with a variant of interest, performing PheWAS on a known disease-associated SNP can identify potential co-morbid conditions,(Byun et al. 2021) as well as traits that may mediate the SNP-disease association.(Pettit et al. 2021) For example, a PheWAS of the lung cancer risk-variant rs16969968 reveals numerous strong associations with various smoking-related behaviors (*e*.*g*., cigarettes per day) that mediate the variant’s association with lung cancer risk.(Millard et al. 2019) Identifying such “intermediate traits” connecting LOAD risk alleles to causal mechanisms of LOAD pathogenesis has clear relevance for disease prevention and population-based risk reduction.(Semmes, Zhang, and Walsh 2018) Furthermore, by identifying traits that have overlapping genetic architecture with a disease of interest, investigators can curate the genetic determinants of such traits to generate an empirical list of candidate-SNPs and test these for pleiotropic association with the disease under study.(Semmes et al. 2020)

The strongest genetic risk factor for sporadic LOAD is the APOE ε4 allele, and a PheWAS of *APOE* gene alleles (ε4, ε3 and ε2) was recently performed.(Lumsden et al. 2020) Although that study identified 14 non-neurologic diseases associated with *APOE* alleles, including hypercholesterolemia and ischemic heart disease, a comprehensive PheWAS analysis of all known LOAD risk loci has not been conducted. We recently published an analytic strategy that integrates GWAS and PheWAS data for a disease of interest in order to identify both epidemiologic traits and trait-associated genetic variants that influence disease risk.(Semmes et al. 2020) By applying our approach to LOAD, we attempt to leverage genetic pleiotropy between LOAD and other traits to identify new LOAD risk factors and to nominate biological pathways through which LOAD risk alleles may exert their pathogenic effects.

## MATERIALS AND METHODS

### Selection of LOAD risk alleles

Hundreds of studies have investigated the genetic epidemiology of LOAD, with associated alleles replicating to highly variable degrees in subsequent research. Associated variants discovered by GWAS are historically more likely to replicate in future studies due to their use of more stringent thresholds for determining statistical significance, lack of an *a priori* hypothesis, and tendency to externally replicate associated variants within the primary publication.(Walsh et al. 2013) We compiled a list of common (minor allele frequency ≥0.01 in European-ancestry populations) LOAD-associated genetic variants for PheWAS analysis from the most recent and largest GWAS meta-analysis of LOAD published at the time, which included 35,274 clinical and autopsy-documented LOAD cases and 59,163 controls genotyped or imputed for 9,456,058 SNPs.(Kunkle et al. 2019) Using a list of LOAD-associated variants reaching canonical genome-wide statistical significance (*i*.*e. P<*5.0×10^−8^) in their study, we performed linkage disequilibrium (LD) pruning (R^2^≤0.15 in European-ancestry populations) using LDlink and retained the variant with the lowest P-value.(Machiela and Chanock 2015) A total of 24 independent LOAD-associated variants were carried forward for PheWAS analyses, including 19 SNPs identified by prior GWAS of LOAD and 5 novel variants that were successfully replicated within that publication.

### Control SNP Set

We compiled a comparison SNP-set to serve as controls for PheWAS analyses. A set of unlinked control SNPs (1000 Genomes Project) was generated using SNPsnap (Broad Institute).(Pers, Timshel, and Hirschhorn 2015) Four control SNPs were matched to the LOAD-associated SNPs on: minor allele frequency (±5%), surrounding gene density (±50%), distance to nearest gene (±50%) and, as a proxy for haplotype block size, the number of other SNPs in LD at R^2^≥0.50 (±50%). For several LOAD risk SNPs, we could not generate more than 4 control SNPs without loosening our matching parameters, but the gain in statistical power achieved beyond a case-to-control ratio of 1:4 is typically minimal.(Hennessy et al. 1999) For one LOAD-associated SNP in the MHC class II region on chromosome 6 (rs9270816), no appropriate control SNPs could be matched. Therefore, rs9270816 was excluded from downstream analyses and a total of 23 LOAD-associated SNPs and 92 control SNPs were carried forward for PheWAS analyses.

### PheWAS analyses

The UK Biobank atlas of genetic associations (http://geneatlas.roslin.ed.au.uk/) was constructed by genotyping 452,264 European-ancestry individuals for 805,426 genetic variants, performing genome-wide SNP imputation and quality-controls, and linking genetic data to EHR data. GeneATLAS contains data for 778 traits (118 quantitative, 660 binary) and associations with 9,113,133 genetic variants (genotyped or imputed). GeneATLAS is searchable and can be queried for genetic (*e*.*g*., SNPs) or phenotypic (*e*.*g*., height) data to assess genotype-phenotype associations.(Canela-Xandri, Rawlik, and Tenesa 2018)

We queried GeneATLAS for trait associations with 23 known LOAD risk SNPs and the control SNP-set (92 matched SNPsnap controls). Summary statistics for traits associated with each queried variant were downloaded from GeneATLAS for downstream analyses. Nominally significant SNP-trait associations (P<0.01) were carried forward to trait-enrichment analyses, as previously described.(Semmes et al. 2020) Although a more stringent p-value threshold for carrying SNP-trait associations forward was considered (*e*.*g*. 0.05/778), this was determined to be too conservative because many of the 778 traits in GeneATLAS have high genetic correlations with each other (*e*.*g*., weight and hip circumference, 0.909; reticulocyte percentage and reticulocyte count, 0.952). Additionally, these individual SNP-trait associations were carried forward for trait-enrichment comparisons between LOAD-associated SNPs and the control SNP-set, and as such the PheWAS significance threshold is somewhat arbitrary so long as it is uniform across all SNP-sets.

PheWAS associations of the 23 LOAD SNPs with 778 phenotypic traits were compared to PheWAS results for the control SNP-set using the Python Programming Environment (https://www.python.org, version 3.6.9). For traits associated with >1 LOAD SNP, we used Fisher’s exact tests to compare the frequency at which individual traits were associated with LOAD SNPs versus control SNPs to determine if traits were enriched for association with known LOAD risk variants. Fisher’s exact p-values for trait enrichment underwent FDR correction to adjust for multiple testing.

### SNPs associated with PheWAS-identified trait (platelet distribution width)

We identified 546 unlinked (R^2^≤0.15 in European-ancestry populations) SNPs with minor allele frequency (MAF) >0.01 that were independently associated with platelet distribution width (PDW) at genome-wide statistical significance in a recent GWAS (N=563,085 subjects).(Vuckovic et al. 2020) Variants, effect alleles, effect sizes, and standard errors were extracted for downstream single-SNP association testing with LOAD and for two-sample Mendelian randomization analysis with LOAD.

### Mendelian randomization analyses

To assess for a causal relationship between PDW and LOAD risk, we performed formal two-sample MR analyses with the R package “MendelianRandomization”.(Yavorska and Burgess 2017; Burgess and Thompson 2013) Using summary statistics of SNP-exposure (*i*.*e*., PDW) and SNP-outcome (*i*.*e*., LOAD) associations, we used the (1) inverse-variance weighted (IVW), (2) MR-Egger, and (3) weighted median methods to test for a causal relationship between PDW and LOAD risk.(Bowden et al. 2016; Bowden, Davey Smith, and Burgess 2015)

### LOAD case-control data

To determine whether PDW-associated SNPs impact risk of LOAD at the single-variant level or under a Mendelian randomization framework, we leveraged SNP summary-level data from a recent GWAS of LOAD. This GWAS included 17,008 European-ancestry LOAD patients and 37,154 controls from the International Genomics of Alzheimer’s Project (I-GAP).(Lambert et al. 2013) Genotypes were imputed with the use of European population reference (EUR) haplotype data from the 1000 Genomes Project, and SNPs that did not pass study-level genotyping quality-control (QC) measures or had a MAF <0.01 were excluded, as were SNPs that did not pass study-level imputation QC measures (INFO score ≥0.30) or were not successfully genotyped/imputed in at least 40% of subjects across studies. The association of LOAD with genotype dosage was analyzed using logistic regression, adjusting for age, sex and ancestry-informative principal components (for three studies with incident LOAD data, Cox proportional hazards models were used). A total of 7,055,881 SNPs with MAF>0.01 are included in the final I-GAP meta-analysis results. Of 546 PDW-associated SNPs with MAF >0.01 from Vuckovic, *et al*.,(Vuckovic et al. 2020) 384 had data available from I-GAP after QC filtering.

### eQTL and *in silico* SNP functional analyses

We characterized SNPs in novel LOAD risk loci using HaploReg to annotate chromatin state and regulatory motifs.(Ward and Kellis 2016) We examined whether variants were protein-binding, located in DNAse hypersensitive sites, promoter or enhancer histone marks, predicted to change transcription factor binding motifs, or functioned as expression quantitative trait loci (eQTL) in diverse tissues,(Ward and Kellis 2016) including the brain.(Zou et al. 2012)

## RESULTS

An overview of the published methodology adapted to our study of LOAD is displayed in **Figure 1**.(Semmes et al. 2020) We first identified 24 common LOAD-associated SNPs from prior GWAS of European-ancestry populations, of which 23 were successfully matched to 4 control SNPs each (**Table 1 and Supplementary Table 1**). PheWAS analyses were then performed with the UK Biobank GeneATLAS database to test each LOAD-associated variant and control variant for association with 778 traits. After determining which PheWAS-identified traits were enriched for association with the LOAD SNP-set (n = 23) compared to the control SNP-set (n = 92), we identified SNPs associated with the enriched PheWAS-identified trait in prior GWAS. Using MR and candidate-SNP approaches, we tested whether enriched traits or trait-associated SNPs conferred LOAD risk in I-GAP data.

**Figure 1.**
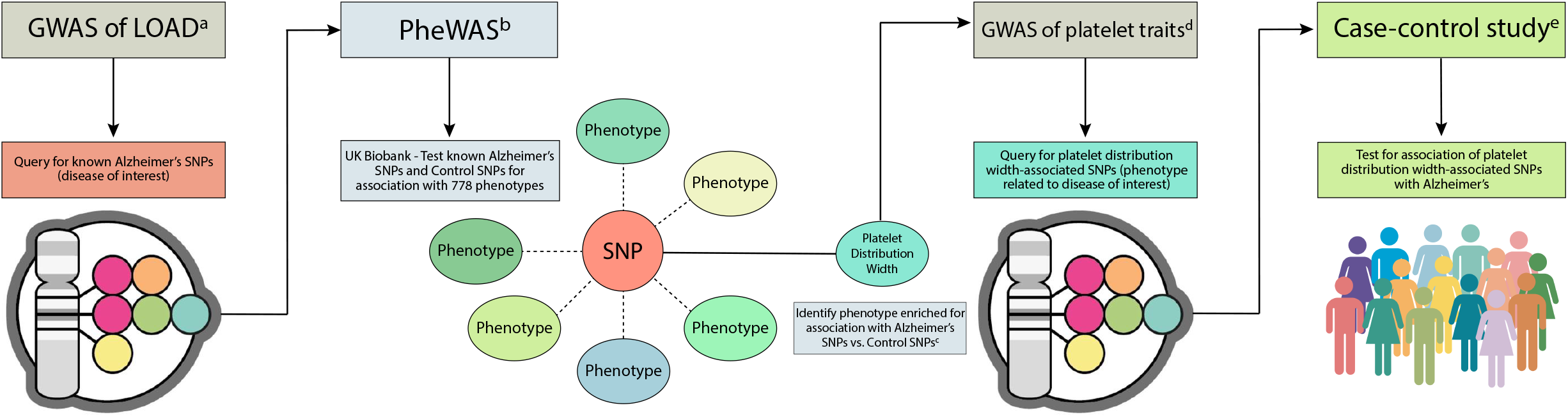
Methodology for integrative analysis of GWAS and PheWAS data. This figure illustrates our approach for investigating phenotype associations with known disease risk-variants in order to identify novel candidate risk loci and/or intermediate phenotypes for subsequent analysis in case-control cohorts. Applying this approach to late-onset Alzheimer’s disease (LOAD) identified platelet distribution width (PDW) as a phenotype enriched for association with LOAD GWAS hits and informed downstream analysis of the role of PDW-associated variants in conferring LOAD risk in a case-control sample. Created with Biorender.com. ^a^ Genome-wide meta-analysis of 35,274 clinical and autopsy-documented LOAD cases and 59,163 controls (Kunkle, *et al*.) ^b^ PheWAS catalog (UK Biobank GeneATLAS) - http://geneatlas.roslin.ed.ac.uk/phewas/ ^c^ Four control SNPs were matched to each LOAD-associated SNP on minor allele frequency (±5%), surrounding gene density (±50%), distance to nearest gene (±50%) and the number of other SNPs in LD at R^2^≥0.50 (±50%) using the SNPsnap software (Broad Institute) - https://data.broadinstitute.org/mpg/snpsnap/ ^d^ Genome-wide meta-analysis of platelet traits in 563,085 subjects (Vuckovic, *et al*.) ^e^ Summary-level SNP association data available from the I-GAP genome-wide meta-analysis LOAD patients and controls (Lambert, *et al*.) Abbreviations: GWAS, genome-wide association study; PheWAS, phenome-wide association study; LOAD, late-onset Alzheimer’s disease; SNPs, single nucleotide polymorphisms.

**Table 1.** Common genetic variants associated with late-onset Alzheimer’s disease (LOAD) in prior GWAS and included in our PheWAS-based enrichment analyses.

| Position (Chr:BP)<sup>a</sup> | rsID | MAF<sup>b</sup> | Function | Nearest Gene |
| --- | --- | --- | --- | --- |
| 1:207802552 | rs4844610 | 0.19 | intronic | *CR1* |
| 2:127892810 | rs6733839 | 0.40 | intergenic | *BIN1* |
| 2:233981912 | rs10933431 | 0.22 | intergenic | *INPP5D* |
| 6:47431284 | rs9473117 | 0.27 | intergenic | *CD2AP* |
| 7:100091795 | rs12539172 | 0.32 | 3' UTR | *NYAP1* |
| 7:143099133 | rs10808026 | 0.23 | intronic | *EPHA1* |
| 8:27219987 | rs73223431 | 0.35 | intronic | *PTK2B* |
| 8:27467686 | rs9331896 | 0.40 | intronic | *CLU* |
| 10:11720308 | rs7920721 | 0.36 | intergenic | *ECHDC3* |
| 11:47380340 | rs3740688 | 0.46 | intronic | *SPI1* |
| 11:59936926 | rs7933202 | 0.40 | intergenic | *MS4A6A* |
| 11:85868640 | rs3851179 | 0.38 | intergenic | *PICALM* |
| 11:121435587 | rs11218343 | 0.04 | intronic | *SORL1* |
| 14:53391680 | rs17125924 | 0.08 | intronic | *FERMT2* |
| 14:92932828 | rs12881735 | 0.22 | intronic | *SLC24A4* |
| 15:59045774 | rs593742 | 0.31 | intergenic | *ADAM10* |
| 16:19808163 | rs7185636 | 0.15 | intronic | *IQCK* |
| 16:79355857 | rs62039712 | 0.10 | intergenic | *WWOX* |
| 17:61538148 | rs138190086 | 0.01 | intergenic | *ACE* |
| 19:1056492 | rs3752246 | 0.18 | missense | *ABCA7* |
| 19:45411941 | rs429358 | 0.14 | missense | *APOE* |
| 20:54997568 | rs6024870 | 0.08 | intronic | *CASS4* |
| 21:28156856 | rs2830500 | 0.30 | intergenic | *ADAMTS1* |

<sup>a</sup>Position in GRCh37/hg19
<sup>b</sup>Minor Allele Frequency in European-ancestry subjects from the 1000 Genomes Project

### PheWAS analyses

216 of the 778 traits in the GeneATLAS database were nominally associated (P<0.01) with >1 of the 23 LOAD-associated SNPs and were tested for enrichment versus the control SNP-set (**Table S2**). A total of eight traits were more likely to be associated with LOAD SNPs than with control SNPs at nominal statistical significance (P<0.05), including: platelet distribution width (PDW), platelet crit, platelet count, leukocyte count, immature reticulocyte fraction, cheese intake, high light scatter reticulocyte percentage, and current tobacco smoking (**Table 2**). PDW was the PheWAS-identified trait most enriched for association with LOAD SNPs, as 11 out of 23 (47.8%) LOAD SNPs were associated with platelet distribution width at P<0.01 compared to 6 of 92 (6.5%) control SNPs (Fisher’s exact P=1.2×10^−5^). Moreover, this association survived both FDR correction (P_FDR_=2.6×10^−3^) and Bonferroni correction for all 778 traits examined (P=9.3×10^−3^). We did not observe any consistent direction of effect between PDW and LOAD risk, as five LOAD risk SNPs were associated with increased PDW and six LOAD risk SNPs were associated with decreased PDW. Although the other seven traits appearing in Table 2 did not demonstrate significant enrichment for association with LOAD SNPs after FDR correction, it is noteworthy that they include two additional platelet-related traits (platelet crit and platelet count).

**Table 2:** Traits more likely to be associated with late-onset Alzheimer’s disease (LOAD) risk SNPs than with control SNPs in PheWAS enrichment analyses.

| Trait | Associated LOAD SNPs | Associated control SNPs | P-value | P-value<sub>(FDR)</sub> |
| --- | --- | --- | --- | --- |
| Platelet distribution width | 11/23 | 6/92 | $1.2 \times 10^{-5}$ | $2.6 \times 10^{-3}$ |
| Platelet crit | 10/23 | 11/92 | $1.4 \times 10^{-3}$ | 0.15 |
| Leukocyte count | 7/23 | 5/92 | $2.2 \times 10^{-3}$ | 0.16 |
| Cheese intake | 4/23 | 1/92 | $5.5 \times 10^{-3}$ | 0.25 |
| Platelet count | 8/23 | 9/92 | $5.9 \times 10^{-3}$ | 0.25 |
| Immature reticulocyte fraction | 5/23 | 3/92 | $7.9 \times 10^{-3}$ | 0.28 |
| High light scatter reticulocyte percentage | 6/23 | 6/92 | 0.014 | 0.39 |
| Current tobacco smoking | 4/23 | 2/92 | 0.014 | 0.39 |

### Mendelian randomization (MR) analyses

Although the eleven LOAD SNPs associated with PDW did not demonstrate associations in a consistent direction (*i*.*e*., increased versus decreased PDW), we further evaluated the relationship between PDW and LOAD risk using a larger set of variants underlying PDW biology. To test for a causal relationship between PDW and LOAD, we used several MR analytical approaches leveraging 384 known PDW-associated SNPs from prior GWAS as instrumental variables and summary statistics for these SNPs from the I-GAP GWAS of LOAD. Estimates from IVW (P_IVW_=0.27), MR-Egger (P_MR-Egger_=0.69, P_MR-intercept_=0.19), and median-weighted (P_MR-median_=0.64) methods were all non-significant, indicating that PDW does not mediate LOAD risk and that there is likely no causal relationship between the traits.

### PDW-associated SNPs as candidate LOAD risk loci

To examine whether individual PDW-associated variants might have pleiotropic effects on LOAD risk, we evaluated single-SNP associations of 384 PDW-associated SNPs with LOAD risk in 17,008 LOAD cases and 37,154 controls from I-GAP. Thirty-six SNPs were nominally associated (P<0.05) with LOAD case-control status (**Table S3**), of which five remained associated after FDR correction (**Table 3**). These associations confirmed known LOAD risk loci near *PICALM, CD2AP, SPI1*, and *NDUFAF6*, and identified a novel risk locus at rs12535226 in the epidermal growth factor receptor (*EGFR*) gene. The rs12535226 variant is located the first intron of *EGFR* and has a minor allele frequency of 48% in European-ancestry populations. *In silico* functional analysis provided strong evidence of this variant having a regulatory role, as it is located in an enhancer region enriched for H3K27Ac histone marks in both brain tissues and neurosphere cultures,(Ward and Kellis 2016) and was identified as an *EGFR* expression quantitative trait locus (eQTL) in both the cerebellum and temporal cortex.(Zou et al. 2012)

**Table 3:** Variants achieving genome-wide statistical significance in a prior GWAS of platelet distribution width (Vuckovic, *et al*.) and their association with late-onset Alzheimer’s disease (LOAD) risk (17,008 patients; 37,154 controls). A total of 384 sentinel variants from Vuckovic, et al. were assessed for association with LOAD risk and appear in Supplementary Table 3, of which five remained associated after FDR correction and appear here.

| Variant ID | Position (Chr:BP)<sup>a</sup> | P-value | P-value<sub>(FDR)</sub> | Nearest gene(s) | Previously-identified LOAD risk locus |
| --- | --- | --- | --- | --- | --- |
| rs3851179 | 11:85868640 | 2.8x10<sup>-15</sup> | 1.1x10<sup>-12</sup> | *EED, PICALM* | Yes |
| rs4711890 | 6:47606712 | 4.9x10<sup>-6</sup> | 9.4x10<sup>-4</sup> | *CD2AP* | Yes |
| rs7107356 | 11:47676170 | 8.4x10<sup>-6</sup> | 1.1x10<sup>-3</sup> | *MTCH2, AGBL2* | Yes |
| rs12535226 | 7:55156419 | 4.3x10<sup>-4</sup> | 0.034 | *EGFR* | No |
| rs4735333 | 8:95955074 | 4.5x10<sup>-4</sup> | 0.034 | *TP53INP1, NDUFAF6* | Yes<sup>b</sup> |

<sup>a</sup>Position in GRCh37/hg19
<sup>b</sup>This region approached genome-wide statistical significance in a prior GWAS of LOAD(Kunkle et al. 2019) (lead SNP rs4735340, P=9.2x10<sup>-8</sup>), but was not independently replicated and thus is not yet considered a validated LOAD risk locus.

## DISCUSSION

We apply an innovative and previously-validated framework for leveraging existing GWAS and PheWAS data to uncover traits associated with Alzheimer’s risk and to identify trait-associated variants as candidate LOAD risk loci. We first curated SNPs associated with LOAD from large GWAS meta-analyses and matched these LOAD risk variants to control SNPs on multiple parameters, including MAF, gene density, distance to nearest gene, and the number of other SNPs in LD. Performing PheWAS on these LOAD and control SNPs using UK Biobank data, we identified platelet distribution width (PDW) as being significantly enriched for association with LOAD risk loci. Although MR analysis did not implicate PDW as having a causal role in LOAD pathogenesis, a number of PDW-associated SNPs demonstrated pleiotropic associations with LOAD risk, including a novel association between LOAD and an *EGFR* eQTL variant.

Although PDW was significantly enriched for association with LOAD risk SNPs, MR analyses did not suggest the presence of vertical pleiotropy, wherein PDW directly contributes to LOAD risk. However, the large number of pleiotropic loci observed to influence both PDW and LOAD suggest that substantial horizontal pleiotropy exists between these traits, which occurs when a variant affects LOAD directly or through multiple pathways not limited to platelet morphology. This appears reasonable given the emerging role of immune function in LOAD pathogenesis,(Lewcock et al. 2020) and is supported by our single-SNP association results that identified known and novel LOAD risk loci using PDW-associated variants as empirically-derived candidate SNPs. Confirming the validity of this approach, our analyses re-identified variants in *SPI1*, a transcriptional activator of myeloid and B cell developmental gene expression previously linked to LOAD pathogenesis. They also identified a PDW-associated variant upstream of *NDUFAF6*, a region that has approached genome-wide statistical significance in a prior GWAS of LOAD (P=9.2×10^−8^) but has not yet emerged as a confirmed risk locus and was therefore not included among the LOAD SNPs used in our PheWAS analysis.

A novel LOAD risk locus in *EGFR* was implicated by our integrated analyses and the risk allele has been associated with elevated EGFR expression levels in human brain tissues.(Zou et al. 2012) EGFR, a member of the Human Epidermal Growth Factor Receptor 1 (ErbB) family, is a transmembrane tyrosine kinase receptor that initiates cell differentiation and proliferation. EGFR expression increases during neurogenesis and gliogenesis, and it activates pathways involved in oncogene expression.(Voldborg et al. 1997) EGFR plays a crucial role in the development of cancer, especially non-small lung cell carcinoma and glioblastoma, and extensive research has been completed evaluating it as a therapeutic target in the oncology setting.(da Cunha Santos, Shepherd, and Tsao 2011; Congdon et al. 2014) However, its function in the pathology of neurodegenerative diseases, such as LOAD, has received comparatively little attention.(Mansour et al. 2021)

A strongly supported theory for LOAD pathogenesis is the amyloid cascade hypothesis, positing that Aβ plaque accumulation leads to “synaptic dysfunction, tau pathology, and neuronal loss”.(Saraceno et al. 2013) The Aβ42 isoform increases in quantity in forms of AD, forming the characteristic amyloid plaques associated with Alzheimer’s neuropathology.(Schmidt et al. 2009) In *Drosophila* model systems, research suggests that Aβ-induced memory loss results from activation of EGFR by Aβ42 oligomers, providing evidence that EGFR may function as a cell membrane receptor of Aβ peptides.(Wang et al. 2012) Following brain injury, EGFR expression increases, leading to the conversion of quiescent astrocytes to reactive astrocytes.(Liu and Neufeld 2007) While quiescent astrocytes have a constructive function in the developing CNS, reactive astrocytes are destructive and lead to neuronal loss. Recent tissue-based analyses have also identified significant differences in EGFR expression and protein levels between LOAD patients and control subjects,(He et al. 2021) and single-cell sequencing analyses implicate EGFR and MAP4K4 upregulation as a biomarker of altered ligand-receptor axis communication between neurons and microglia in AD patient brains.(Jian et al. 2021) These emerging data complement our findings and lend support to ongoing work that seeks to repurpose EGFR-targeted cancer therapies to inhibit EGFR as a potential pathway to rescuing memory loss and preventing neuronal loss in LOAD patients.(Tavassoly, Sato, and Tavassoly 2020)

There are several limitations to our study and valid concerns of our hybrid GWAS-PheWAS approach. For the PheWAS analyses, we used a p-value threshold of <0.01 to carry a SNP-trait association forward to enrichment analyses, rather than a Bonferroni-corrected threshold (*i*.*e*., 0.05/778 traits). However, we found that this threshold maximized study power in our previous application of this GWAS-PheWAS approach to leukemia etiology.(Semmes et al. 2020) Additionally, this threshold is only for carrying traits forward to subsequent enrichment analyses, and associations from these enrichment comparisons were subjected to FDR correction to account for the number of traits evaluated for potential enrichment with LOAD SNPs. Another limitation is that our analysis relied upon traits evaluable in the U.K. Biobank, and therefore does not permit us to assess enrichment for potential disease-related traits that may have relevance to LOAD pathology, such as cerebral microbleeds or history of viral infections. Our PheWAS analyses also relied upon well-validated LOAD risk loci that had reached genome-wide statistical significance and been replicated in external datasets. Inclusion of additional putative risk alleles meeting less stringent cutoffs could potentially improve study power, but may also dilute effects by including false-positive LOAD associations. Since completing our analyses, an additional GWAS meta-analysis of LOAD has been published, identifying several additional risk alleles.(Wightman et al. 2021) However, more than 50% of LOAD cases included in this study were “proxy cases” with a positive family history of LOAD, which may increase statistical power at the expense of precision. Such studies are useful adjuncts to study LOAD etiology, but do not lend themselves to MR approaches due to potential misspecification of SNP effect sizes.

Although multiple GWAS in the past decade have contributed to our understanding of inherited susceptibility to LOAD, there remains significant missing heritability.(Wightman et al. 2021; Kunkle et al. 2019; Lambert et al. 2013) A recent review on the benefits and pitfalls GWAS emphasized the need for novel analytic approaches to enhance our understanding of genotype-phenotype associations in the post-GWAS era and the utility of large biorepository databases linking EHR and genotyping data, polygenic scores, and innovative study designs.(Wang, Li, and Hakonarson 2010) While increasing GWAS sample size may reveal more associations, new methods for analyzing the wealth of existing data are essential, particularly in unraveling the underlying biological pathways involved in SNP-disease associations. In summary, by integrating GWAS and PheWAS datasets to decipher the complex etiology of LOAD, we identify horizontal pleiotropy with PDW and implicate EGFR variation in disease predisposition. These results should stimulate further mechanistic studies on the role of EGFR in AD pathogenesis to dissect their potential relationship. Such findings will have utility in informing the rational design of future prevention and treatment strategies for an aging population.

## Data Availability

This study leverages publicly available human data, which can be obtained from UK Biobank and the I-GAP Consortium.

http://geneatlas.roslin.ed.ac.uk/

https://www.niagads.org/datasets/ng00036

